# Deep Learning Enhances Detection of Extracapsular Extension in Prostate Cancer from mpMRI of 1001 Patients

**DOI:** 10.1101/2024.05.21.24307691

**Authors:** Pegah Khosravi, Shady Saikali, Abolfazl Alipour, Saber Mohammadi, Max Boger, Dalanda M. Diallo, Christopher Smith, Marcio Covas Moschovas, Iman Hajirasouliha, Andrew J. Hung, Srirama S. Venkataraman, Vipul Patel

**Author notes:** Corresponding authors: Correspondence to be sent to Department of Biological Sciences, New York City College of Technology, The City University of New York, 285 Jay Street, Brooklyn, NY 11201, USA. These authors are co-first authors (Pegah Khosravi, Shady Saikali).

## Abstract

Extracapsular extension (ECE) is detected in approximately one-third of newly diagnosed prostate cancer (PCa) cases at stage T3a or higher and is associated with increased rates of positive surgical margins and early biochemical recurrence following radical prostatectomy (RP). This study presents the development of AutoRadAI, an end-to-end, user-friendly artificial intelligence (AI) pipeline designed for the identification of ECE in PCa through the analysis of multiparametric MRI (mpMRI) fused with prostate histopathology. The dataset consists of 1001 patients, including 510 pathology-confirmed positive ECE cases and 491 negative ECE cases. AutoRadAI integrates comprehensive preprocessing followed by a sequence of two novel deep learning (DL) algorithms within a multi-convolutional neural network (multi-CNN) strategy. The pipeline exhibited strong performance during its evaluation. In the blind testing phase, AutoRadAI achieved an area under the curve (AUC) of 0.92 for assessing image quality and 0.88 for detecting the presence of ECE in individual patients. Additionally, AutoRadAI is implemented as a user-friendly web application, making it ideally suited for clinical applications. Its data-driven accuracy offers significant promise as a diagnostic and treatment planning tool. Detailed instructions and the full pipeline are available at https://autoradai.anvil.app and on our GitHub page at https://github.com/PKhosravi-CityTech/AutoRadAI.

## Introduction

Accurate preoperative staging of prostate cancer (PCa) plays a pivotal role in tailoring optimal treatment strategies, ensuring that patients neither undergo undertreatment nor overtreatment. Among the critical determinants of disease progression, the presence of extracapsular extension (ECE), identified as stage T3a or beyond, emerges as a prominent factor, encompassing approximately one-third of newly diagnosed PCa cases ^1,2^. ECE’s significance lies in its close association with elevated rates of positive surgical margins and early biochemical recurrence following radical prostatectomy (RP), making it a key consideration in clinical decision-making ^3,4^. ECE has a significant prognostic value, decisively marking whether the cancer has infiltrated beyond the confines of the prostate gland. Many studies have proposed that not only the presence but also the amount of ECE is an independent predictive factor for biochemical recurrence (BCR) ^5,6^. This further highlights the relevance of accurately predicting the presence of ECE before surgery.

While predictive nomograms based on clinicopathological attributes as well as MRI have been previously developed and validated ^7,8^, the intricate anatomy of the prostate and the subtle contrast between cancerous and healthy tissues present formidable challenges for ECE detection on magnetic resonance imaging (MRI) images. In the ever-evolving landscape of medical imaging and diagnosis, machine learning (ML) and deep learning (DL) algorithms have emerged as revolutionary tools with widespread applications ^9–12^. Within this context, DL models, a subset of ML, have demonstrated remarkable capabilities in deciphering intricate patterns and relationships within medical images ^13^. This extends to the domain of PCa diagnosis, where DL algorithms have shown promise ^14–17^. Also, with the lack of a truly objective method of assessment to evaluate the presence of PCa on MRI, one must turn to indirect signs to confirm or reject the likelihood of a cancer diagnosis.

Traditionally, identifying ECE as well as lesion identification (PIRADS classification) on MRI images has been a time-intensive process heavily reliant on the expertise of radiologists. However, the advent of DL has opened new avenues for automating this intricate task. By training convolutional neural networks (CNNs) on extensive datasets of annotated MRI scans, these algorithms can learn to recognize subtle visual cues indicative of ECE ^18,19^. The utilization of lightweight CNN architectures, such as the ones proposed in this study, further optimizes the delicate balance between accuracy and efficiency.

As we explore the landscape of current diagnostic technologies and their limitations in detecting ECE, the need for innovative solutions becomes clear. In response to this challenge, we introduce AutoRadAI (**Figure 1**) in this paper. AutoRadAI represents a breakthrough in medical imaging technology: a comprehensive, fully automated pipeline specifically designed for ECE detection. By leveraging the capabilities of two advanced DL models, ProSliceFinder and ExCapNet, AutoRadAI offers a systematic framework for the processing and refinement of images. This process extends from the radiologist’s bench, through a series of sophisticated analytical stages, to the generation of outcomes that are easily interpretable by medical professionals. Our development is not just about technological innovation; it aims to streamline and enhance the diagnostic workflow. Our study leverages the largest cohort to date, encompassing 1001 patients, to predict ECE in PCa, setting a new benchmark in the field. This extensive population size not only enhances the statistical power of our analysis but also underscores the potential for our findings to be generalized across diverse clinical settings. A key component of this enhancement is the development of a web application that utilizes user-friendly, end-to-end pipelines as its core. Designed for clinical use, the application promises to facilitate the rapid and reliable identification of ECE across diverse MRI datasets. By improving both the accuracy and consistency of diagnoses, this new approach seeks to transform patient management. It emphasizes early diagnostic intervention and supports more precise treatment planning, thereby making a significant contribution to the advancement of patient care.

**Figure 1:**
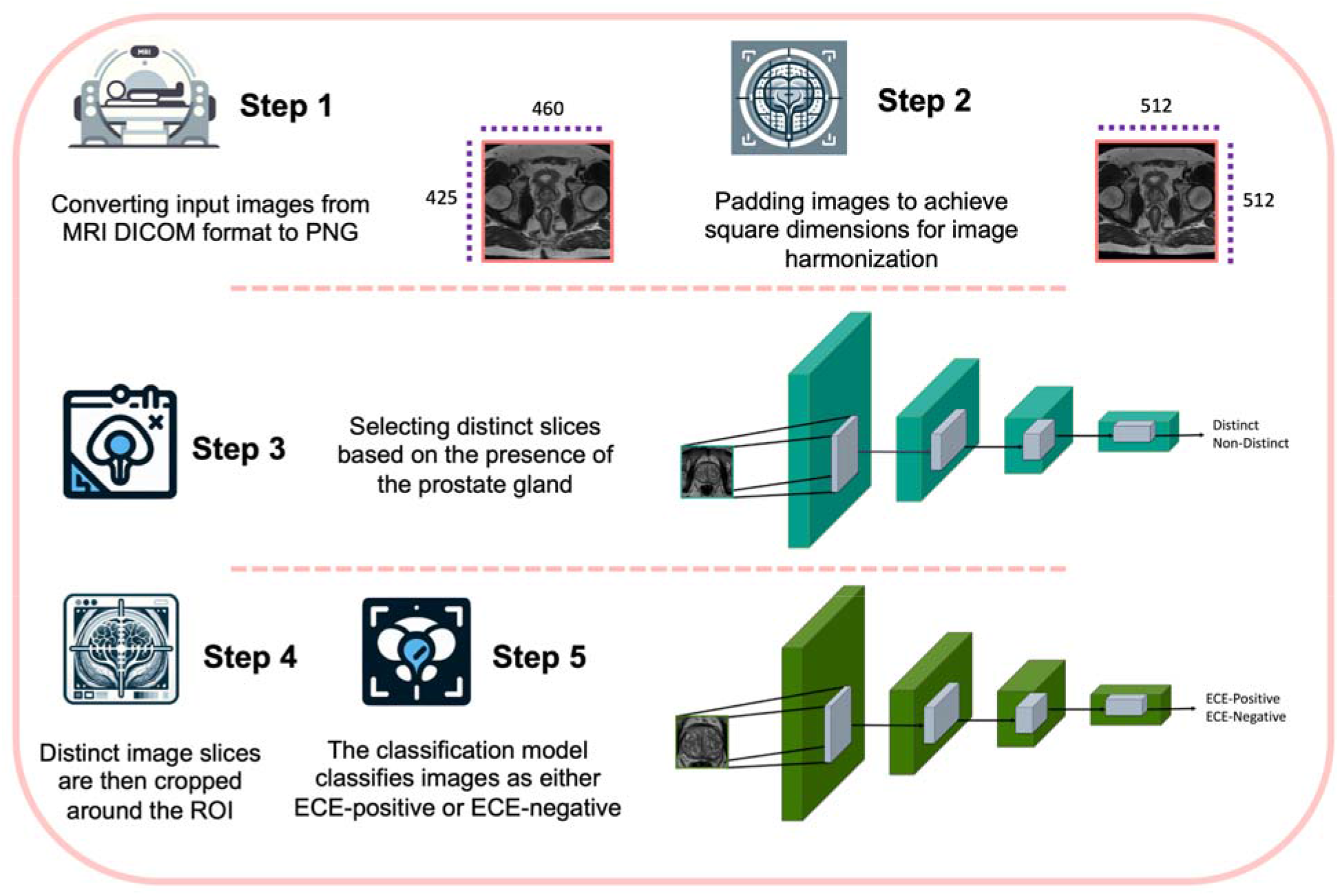
Schematic representation of the AutoRadAI pipeline. This figure illustrates the end-to-end workflow of our proposed system, starting from the initial image acquisition at the radiologist’s bench, through the processing and analysis by multi-CNN (ProSliceFinder and ExCapNet), to the final ECE detection. The output from Step 2 is utilized as the input for training the first deep learning model in Step 3, while the output from Step 4 serves as the input for training the second deep learning model in Step 5.

## Results

### Distinct Scans and ECE Detection Evaluation

Our analysis of the AutoRadAI system, which integrates a sophisticated multi-CNN architecture, underscores its remarkable effectiveness in detecting ECE in preoperative MRI scans of PCa patients. Specifically, **Figure 2** demonstrates the proficiency of the ProSliceFinder and ExCapNet models in identifying relevant scans and accurately predicting ECE. The ProSliceFinder algorithm, designed to analyze individual image slices, exhibited impressive performance metrics on a blind test set comprising 467 MRI slices. It achieved an AUC of 0.92, Sensitivity of 0.91, Specificity of 0.80, and an overall ACC of 0.85, thereby effectively identifying slices depicting the prostate gland using T2W imaging, as illustrated in **Figure 2a. Figure 2a**’s ROC curve and confusion matrix reveal a high degree of diagnostic accuracy. The confusion matrix shows that 183 distinctive slices and 216 non-distinctive slices were correctly identified, underscoring the model’s capability to discriminate between distinctive and non-distinctive scans effectively.

**Figure 2:**
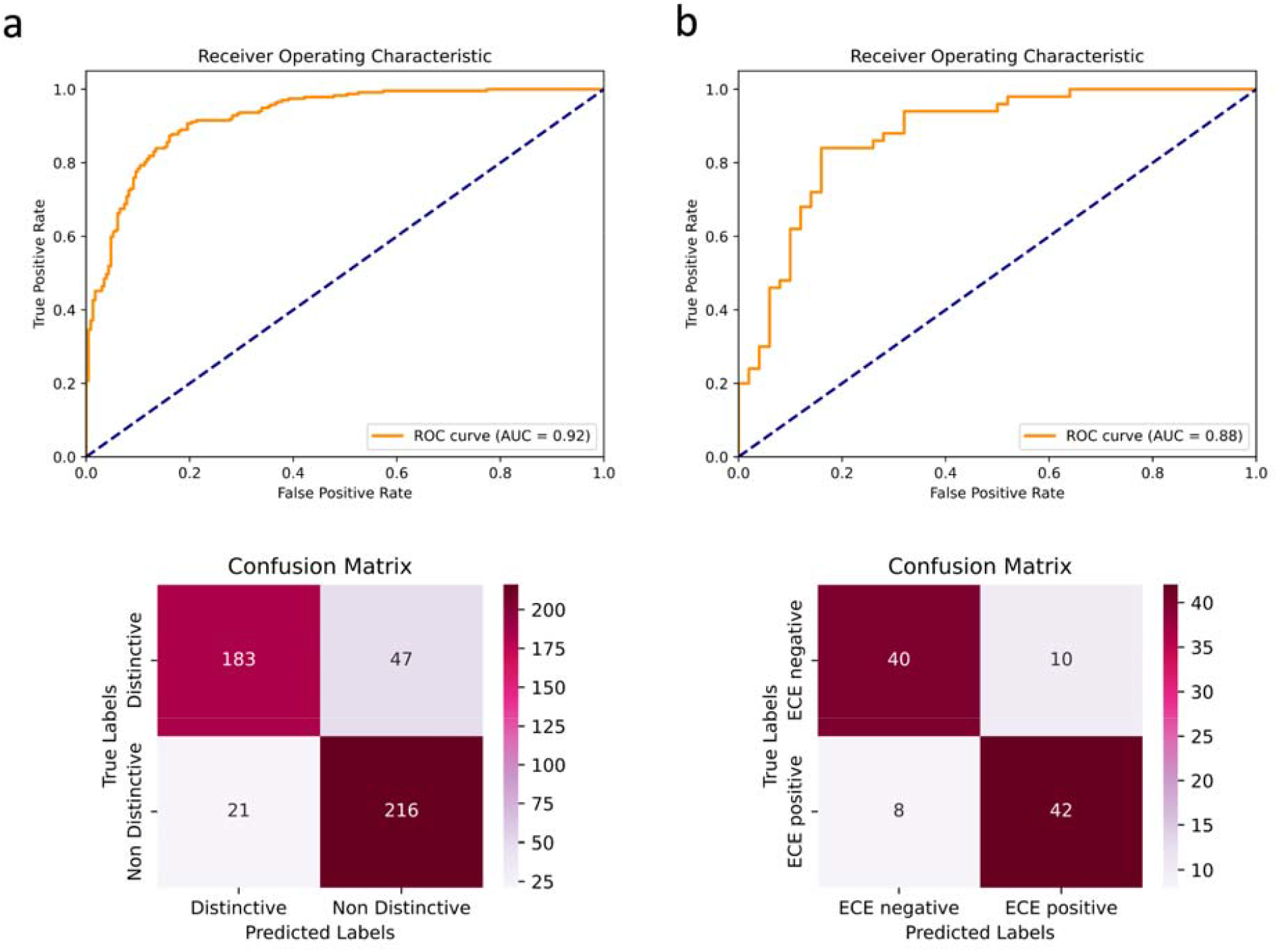
Performance of ProSliceFinder (a) and ExCapNet (b) for detecting Distinct (visible prostate gland) slices and ECE detection, respectively using ROC curves and confusion matrix.

Complementing this, the ExCapNet model, which assesses data at the patient level, garnered an AUC of 0.88, Sensitivity of 0.84, Specificity of 0.80, and ACC of 0.82. This analysis, performed on a blind test set featuring 100 randomly selected patients, is highlighted in **Figure 2b**. The ROC curve and confusion matrix for the ExCapNet model illustrates its capability to effectively classify ECE status with a notable balance between sensitivity and specificity—42 true positives and 40 true negatives. This model capitalizes on the slice-level insights provided by ProSliceFinder to make informed patient-level ECE predictions, thereby supporting its application in clinical settings where accurate ECE assessment is crucial for effective treatment planning.

### Decision Tree Analysis for Predicting Extracapsular Extension

In our investigation, we undertook a comparative analysis of three ML models— Decision Tree, Random Forest, and XGBoost—to assess their efficacy in predicting ECE using the ISUP grade group and patient age across a cohort of 1001 patients. Additionally, we explored the impact of incorporating AI-derived features on the predictive accuracy within a subset of 100 randomly selected test patients. Initially, our dataset, comprising 1001 patients, was analyzed holistically, followed by a nuanced evaluation of two distinct 100-patient groups within our blind test set, differentiated by the integration of AI-derived features.

For the larger cohort of 1001 patients, our findings revealed moderate predictive capabilities across the models, with the Decision Tree achieving an ACC of 0.64 and an AUC of 0.65, Random Forest with an ACC of 0.62 and AUC of 0.65, and XGBoost leading slightly with an ACC of 0.63 and an AUC of 0.68 (illustrated in **Figure 3a**). This initial analysis highlighted XGBoost’s marginal superiority in AUC, suggesting nuanced variations in model performance.

**Figure 3:**
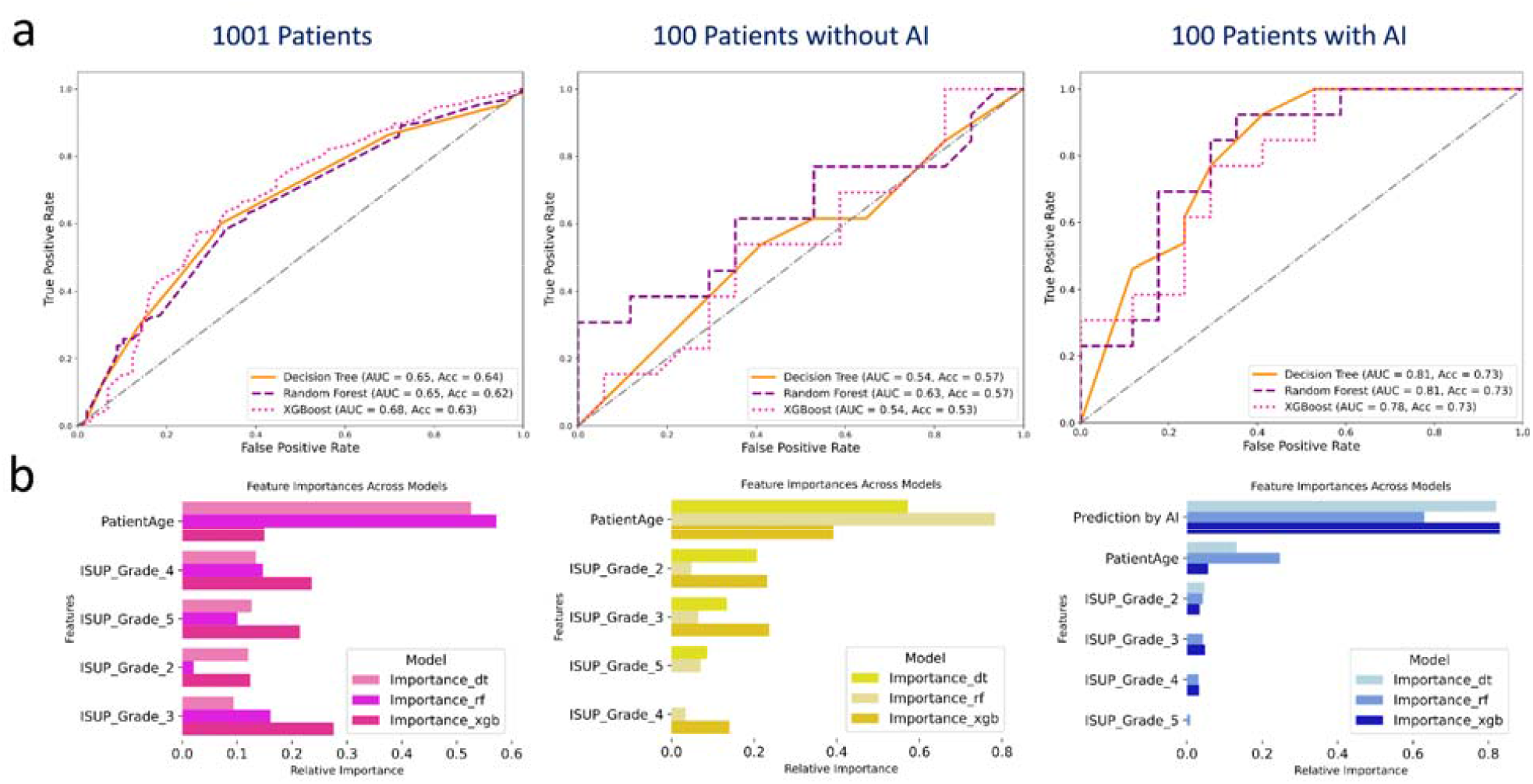
Analytical performance of Decision Tree, Random Forest, and XGBoost models, which incorporate histopathological grades and patient age for predictive analysis. It provides a comparative evaluation of these models’ predictive accuracy in two distinct scenarios: across a dataset of 1001 patients (Panel a) and within a subset of 100 patients, distinguished by the absence (Panel b) and presence (Panel c) of AI-derived features. The lower portion of the figure enumerates the key features determined by each algorithm to be most critical for predictive performance, segmented by model and dataset.

Upon excluding AI-derived features in the subsequent 100-patient cohort (**Figure 3b**), a noticeable decrement in performance metrics was observed across all models, underscoring the significant role AI features play in enhancing predictive accuracy. Conversely, the integration of AI features not only recuperated but also enhanced the performance of the models. Specifically, the Decision Tree, Random Forest, and XGBoost models showed a significant increase, with their AUC values rising to 0.81, 0.81, and 0.78, respectively. Additionally, all three models achieved the same ACC of 0.73. This improvement accentuates the additive value of AI insights to the predictive analytics framework.

Further elucidating the impact of AI integration, **Figure 3c** presents the feature importance rankings, highlighting a pivotal shift toward AI predictions as a significant determinant of ECE, surpassing traditional clinical markers such as patient age and ISUP grade. This evolution in feature importance rankings evidences the potent synergy between clinical attributes and AI-derived features, fostering a refined approach to patient stratification.

The empirical evidence from our analysis strongly advocates for the amalgamation of traditional clinical features with AI-driven insights. This integration not only enriches the predictive landscape but also significantly elevates the performance of ML models within clinical settings, paving the way for more informed and nuanced patient care strategies.

### Comparative Performance: Radiologist Assessment vs. AutoRadAI

In the concluding phase of our study, we tasked three board-certified radiologists with independently assessing the presence of ECE in MRI scans, without prior knowledge of actual pathology outcomes or the existing formal radiology reports. This blind evaluation involved the same cohort of 100 patients previously examined by our AI system, AutoRadAI, aiming to compare the diagnostic accuracy of radiologists and AI against the ground truth from pathology reports (**Figure 4**).

**Figure 4:**
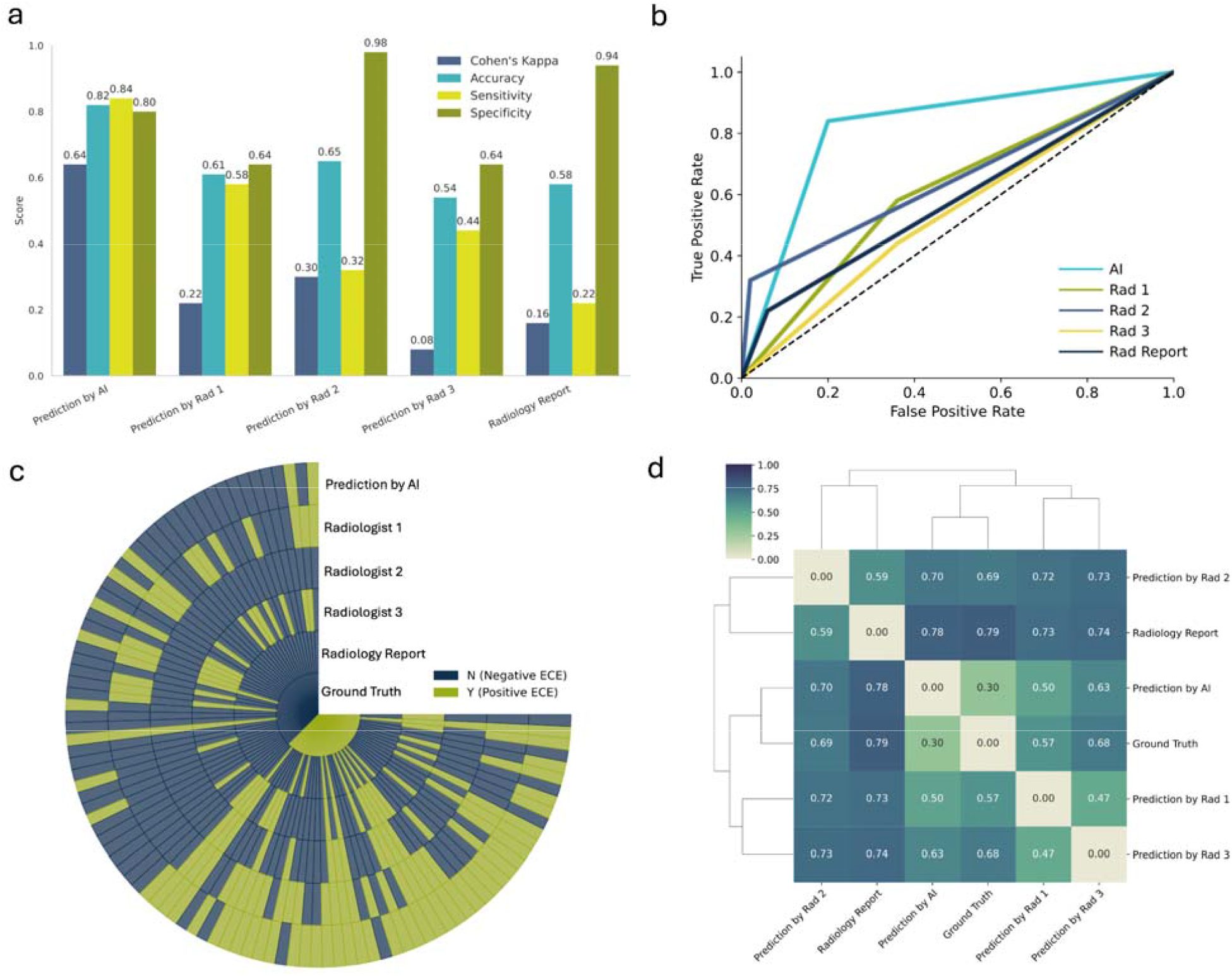
Performance comparison of AutoRadAI for ECE detection in MRI scans against radiologists, including Cohen’s Kappa, Sensitivity, Specificity, and Accuracy metrics (a), AUC (b), Circular heatmap (c), and Dendrogram (d) for AI versus radiologist predictions.

The radiologists’ diagnostic performance quantified using AUC and ACC, recorded values of 0.61, 0.65, and 0.54, respectively, with the formal radiology report yielding an ACC of 0.58 (**Figure 4a and 4b**). These figures suggest that determining ECE status solely through imaging remains challenging, even for seasoned practitioners. In contrast, AutoRadAI demonstrated superior classification capabilities with an ACC of 0.82. Its agreement with the gold-standard pathology results, measured by Cohen’s kappa score, was Substantial (kappa = 0.64), notably higher than that of the individual radiologists (kappa = 0.22, 0.30, and 0.08) and the radiology report (kappa = 0.16), indicating a significant advancement in reliability and accuracy (**Figure 4a**).

To visualize these findings, **Figure 4c** employs a circular heatmap, and **Figure 4d** shows a dendrogram. The heatmap illustrates the degrees of concordance and discordance among the assessments by the radiologists, the formal radiology report, and the confirmed pathology cases, highlighting the spectrum of diagnostic alignment. Meanwhile, the dendrogram, constructed using the Jaccard distance index, clusters the radiologists based on the similarity of their diagnostic decisions and contrasts these with AutoRadAI’s outcomes. Jaccard distances of 0.57, 0.69, 0.68, and 0.79 were observed among the radiologists, with the AI system showing a closer alignment to the ground truth (Jaccard distance = 0.30), revealing distinct diagnostic approaches and accuracy levels (**Figure 4d**).

These analyses, especially the stark contrast in performance between the human experts and AutoRadAI, underline the significant potential of AI to enhance medical diagnostics by providing more reliable and accurate ECE assessments.

## Discussion

The preoperative detection of ECE significantly influences the surgical approach during RARP, directly impacting the postoperative recovery of erectile function. ECE, an adverse pathological sign present in a third of prostate cancer patients at diagnosis ^2^, is associated with increased rates of positive surgical margins and early biochemical recurrence post-prostatectomy. This underscores the critical need for accurate preoperative ECE prediction to tailor surgical plans and ensure safe margin resection without compromising functional outcomes. Despite the validation of clinical nomograms for ECE prediction ^7,8,20,21^, these were developed prior to the widespread adoption of mpMRI, highlighting a gap in leveraging modern imaging in surgical planning.

MRI’s superior sensitivity (>90%) in detecting clinically significant prostate cancer ^22^ offers a rich dataset for surgical planning. Recent advances in ML and DL, particularly in radiomics, have shown promise in enhancing ECE prediction. ^18^ reported that a PAGNet model using a single-slice image yielded a moderate AUC (95% CI, 0.63-0.81), with AI assessments outperforming expert radiologists in external validation datasets. Recent advancements in ML for medical imaging have led to significant achievements in predicting ECE in prostatectomy specimens. ^23^ developed a logistic regression model using a dataset from 139 patients, incorporating clinical, semantic, and radiomic data. This model demonstrated superior predictive performance, with an area under the AUC of 0.93 and an accuracy of 78%. Notably, this model’s performance on the test dataset was superior to that on the training dataset, with an AUC of 0.93 compared to 0.88. This unusual finding suggests an exceptional generalization of the model but also raises questions about the potential overfitting or the representativeness of the test dataset, given the small sample size. This anomaly highlights the need for further validation to ensure the model’s robustness and generalizability. ^24^ utilized a support vector machine (SVM) model trained on a dataset from 193 patients, achieving an accuracy of up to 79% and an AUC of 0.80 on an independent test dataset. The relatively modest performance of the SVM model, despite the larger dataset size, underscores the importance of feature selection and the challenges in generalizing models across diverse patient cohorts. This comparison illustrates the critical balance between dataset size, feature selection, and model complexity, suggesting avenues for future research to enhance model performance and generalizability, especially in the context of limited data availability.

Our study extends these findings by demonstrating the value of multi-slice image analysis through ProSliceFinder, providing a richer dataset and accommodating the variability inherent in MRI scans. Our approach using ExCapNet at the patient level revealed an AUC of 0.88 and ACC of 0.82, markedly surpassing the diagnostic accuracy of radiologists and traditional radiology reports. This suggests the AI model’s proficiency in detecting nuanced cues within MRI slices—cues that may elude even expertly trained eyes. Incorporating texture analysis-based radiomics, as suggested by ^25^, could further refine our ability to distinguish ECE, supporting the potential of a multifaceted AI approach in augmenting diagnostic precision. In our decision tree model, we integrated clinical parameters like Gleason score and age to highlight the synergistic potential of melding clinical factors with AI predictions for detecting ECE. ^26^ demonstrated that the inclusion of key radiomics features alongside clinical parameters (age, PSA, Gleason grade, PIRADS) significantly improves predictive accuracy. This supports the effectiveness of a multimodal approach in clinical diagnostics, aligning with the insights from a recent review study by ^27^. One of the distinguishing features of our investigation is the utilization of the largest study population thus far in the realm of ECE prediction, with 1001 patients. This unprecedented scale not only fortifies the credibility of our findings but also positions our study as a pivotal reference point for future research in accurately predicting ECE in prostate cancer patients.

Nevertheless, our study is not without limitations, including potential biases due to its retrospective nature and the exclusive use of T2W axial images, which, despite being favored by radiologists for ECE detection, may limit the scope of our analysis. We did obtain the patient pool from one single clinical practice, despite the MRI images being obtained from several different machines. We classified the images according to the staging information obtained from the final surgical pathology without reanalyzing the slides. While external validation is a common step in affirming the applicability of diagnostic tools across varied populations, the unique nature of our dataset, primarily sourced from patients presenting to one institution, presents a limitation in this regard. This dataset, however, is not without its strengths. Its heterogeneity is a testament to its comprehensive coverage, encompassing a wide array of patient demographics, clinical presentations, and imaging characteristics. The diversity within our dataset ensures that our findings are not only robust but also generalizable to a broad spectrum of clinical scenarios. External validation remains a future priority to ensure the model’s effectiveness in clinical practice, and expanding the dataset to include a broader range of MRI sequences could further enhance its diagnostic capabilities.

The exceptional performance of our multi-CNN model, AutoRadAI, underscores the transformative potential of DL in the preoperative assessment of PCa. Not only does it facilitate accurate ECE detection, but it also supports nuanced clinical decision-making, impacting treatment strategies and patient outcomes. Looking ahead, expanding external collaborations to validate our findings and incorporating additional imaging sequences could bolster the model’s utility across diverse clinical settings. AutoRadAI represents a significant step forward in the use of AI for ECE detection. By integrating sophisticated DL models with a streamlined image processing framework, we aim to revolutionize the diagnostic workflow, offering a web application that enables quick, reliable ECE identification. This initiative has the potential to improve diagnostic accuracy and consistency, promoting early intervention and precise treatment planning, thereby enhancing patient care.

In examining over a thousand patient journeys, AutoRadAI not only navigates the complexities of prostate cancer diagnosis but also highlights the importance of personalized care, reinforcing the notion that each patient’s story is a critical element of the broader narrative in medical diagnostics. In conclusion, our study introduces an advanced DL model for preoperative ECE detection in MRI scans of prostate cancer patients, showcasing notable accuracy and efficiency. This model, validated with the largest patient cohort for ECE prediction, represents a significant advancement in medical diagnostics, enhancing surgical planning and patient outcomes. While our results are promising, ongoing refinement and external validation are essential to integrate this technology fully into clinical practice. Our findings lay the groundwork for future research, advocating for the incorporation of AI to meet clinical diagnostic needs effectively.

## Methods

### Study Population

After obtaining approval from the relevant institutional review boards, consecutive patients who underwent Robotic-Assisted Radical Prostatectomy (RARP) between January 2018 and November 2023, and had a preoperative multiparametric MRI (mpMRI) of the prostate available for review, were retrospectively identified from our independent, prospectively maintained databases. Patients were classified into two groups based on the extent of their disease: pT2 and pT3, as defined by the American Joint Committee on Cancer (AJCC) staging system. Eligibility criteria for inclusion in this study were delineated as follows: individuals who received surgical treatment at Advent Health/Global Robotics Institute; those who underwent a 3T mpMRI of the prostate, without the use of an endorectal coil, within one year preceding their surgical intervention; and patients with a pathological diagnosis of PCa. The exclusion criteria included patients without preoperative MRI images; those with MRI images older than one year; and patients who received neoadjuvant hormonal treatment, though interventions for benign prostatic hyperplasia or bladder outflow obstruction were deemed acceptable.

MRIs were conducted at multiple centers, including our own, and subsequently uploaded to our picture archiving and communication system (PACS) for storage and data extraction. The imaging equipment at these centers was sourced from a variety of vendors. All surgeries were performed by a robotic surgery expert, and the surgical pathology was evaluated by a genitourinary pathologist with over a decade of specialized experience.

### Data Preparation

To meet our research goals, we carefully curated a comprehensive dataset of preoperative MRI scans from patients with PCa. This dataset was created through a detailed curation and annotation process by fellowship-trained urologists. It includes a vast array of T2-weighted (T2W) MR images, each precisely labeled with corresponding pathology results to denote the presence or absence of ECE, categorized as ECE positive for presence (= 510 patients) and ECE negative for absence (= 491 patients), respectively. The images were saved in PNG format and only the T2W axial image series of the prostate were used.

The dataset consists of 21,706 images from 1,001 patients, sourced from various vendors across several years (2018-2023). These images required standardization to achieve uniform dimensions, ensuring consistency throughout the dataset. Moreover, the images originated from multiple centers, which were later uploaded onto our hospital PACS system before retrieval. This diversity introduces additional complexity to the necessary preprocessing steps. To overcome these challenges, we created two specialized scripts: the first for converting images into a consistent format, and the second for padding the images. This padding ensures the original images are preserved while standardizing their dimensions for subsequent processes. Through this method, we achieve a uniform resolution of 512x512 pixels across all images, maintaining the original resolution and quality of each. Additionally, we implemented a normalization process to further ensure consistency across the dataset (steps 1 and 2 in **Figure 1**).

Building on the findings of ^15^, which demonstrated the effectiveness of the multislice method over single-slice techniques for training CNNs in image analysis, our research adopts a similar strategy. We utilize the ProSliceFinder DL model to process multiple MRI slices per patient automatically. This multislice approach not only increases the volume of data (breadth) but also enhances the granularity of information captured from each patient (detail). By analyzing several contiguous slices, we gain a more comprehensive representation of the anatomical structures, which is crucial for accurate diagnosis and analysis. This method significantly enriches our dataset, enhancing both its breadth and detail, thereby providing a more robust basis for our predictive models. We have carefully curated our primary dataset to include two types of images as classified by expert radiologists: ‘Distinct slices’, which clearly show the prostate gland, and ‘Non-Distinct slices’, which depict the pelvic anatomy, including muscles and other organs, but do not provide a clear view of the prostate gland itself.

The dataset is balanced, consisting of 3,660 images, with 1,828 being Distinct slices and 1,832 Non-Distinct, encompassing a broad range of anatomical details for robust model training. This deliberate selection process significantly enriches our dataset, enabling our CNN model to more effectively discern the complex subtleties and variations present within MRI images. Such an approach markedly increases the robustness and accuracy of our analysis. Consequently, the model automatically selected all the Distinct images from the original dataset which led to building a dataset encompassing a total of 6,162 MRI images for 1001 patients, including 3,181 images indicating the absence of extracapsular extension (ECE-) and 2,981 images confirming its presence (ECE+), with an average of 6.5 slices for ECE-patients and 5.8 slices for ECE+ patients, respectively (step 3 in **Figure 1**).

Prior to classification by ExCapNet into two categories, ECE positive (ECE+) and ECE negative (ECE-), images undergo a critical preprocessing step to ensure optimal focus on the relevant anatomical region. Specifically, it is imperative to crop the images around the prostate gland to ensure the algorithm’s analysis is focused on the presence of the tumor either within or immediately adjacent to the prostate gland or seminal vesicle rather than invading the bladder, rectum, levator ani and obturator internus muscles, and the pelvic bone. To accomplish this targeted image cropping, we developed a specialized script, delineated as step 4 in **Figure 1**.

Upon completion of this preprocessing, the images are aptly prepared for the final analytical phase—classification by ExCapNet (depicted as step 5 in **Figure 1**), which discerns between ECE+ and ECE-cases.

### Multi-CNN Model Characteristics

The essence of our DL approach in this study is rooted in the adoption of a multi-convolutional neural network (multi-CNN) model, which is a critical element of our methodology for detecting ECE in PCa. The multi-CNN model stands out due to several key features that collectively bolster its capability in the complex task of identifying ECE from MRI scans of PCa patients. The underlying structure of the multi-CNN model comprises two sequential CNN algorithms, each designed with specific, interconnected functions in the ECE detection workflow. Notably, both CNNs are trained from scratch, eschewing the use of any pre-trained models in this research. Initially, the ProSliceFinder algorithm is responsible for identifying and selecting the most informative and diagnostically significant MRI slices for each patient. Following this, the ExCapNet algorithm utilizes these chosen slices to categorize patients into positive or negative ECE groups. This classification is based on MRI images that have been labeled in accordance with ground truth post-surgery pathology results.

In our pursuit of a tailored solution for detecting ECE within MRI scans, we opted to train each algorithm from scratch, a strategy that bypasses the limitations tied to the pre-existing knowledge found in transfer learning. This approach grants our models a deeper and more nuanced comprehension of our specific dataset, enabling them to adjust their parameters with enhanced precision for the distinct features of MRI images pertinent to ECE detection. By designing our models specifically for this task, we inherently avoid the common issue of overfitting, which frequently affects complex, pre-trained models when they are repurposed for specialized applications.

Our CNN architectures are designed to be efficient and effective compared to more deeply layered counterparts (**Figure 5a**). This design choice not only minimizes the risk of overfitting but also improves the models’ ability to predict using new, unseen MRI data effectively. Furthermore, the streamlined nature of our models aligns with the practical requirements of clinical environments. They strike a careful balance between computational efficiency and diagnostic precision, making them highly suitable for quick adoption into clinical workflows. The models’ reduced processing demands enable swifter, more efficient analyses without compromising on accuracy, addressing the critical needs of healthcare settings where both time and precision are of utmost importance.

**Figure 5:**
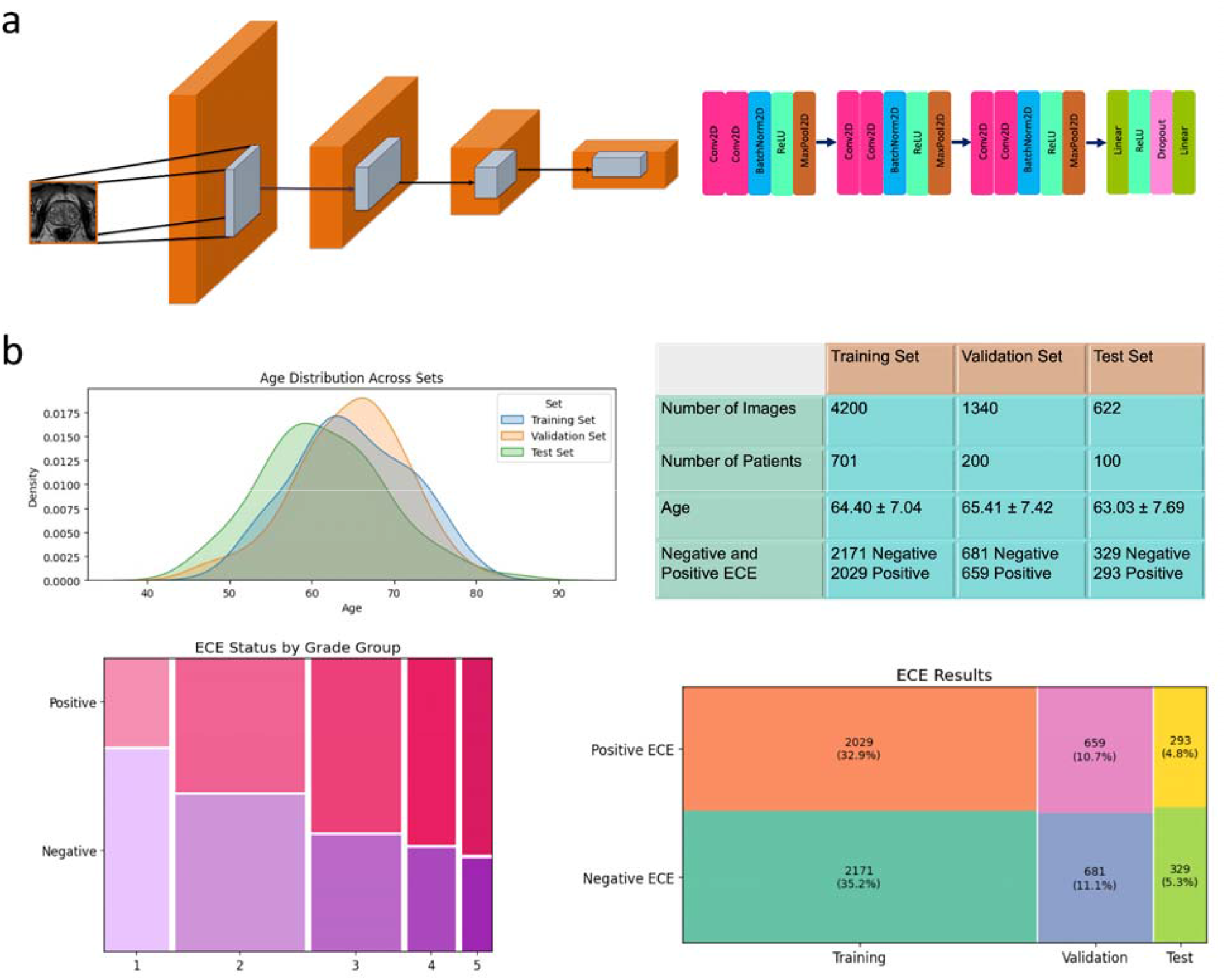
The architectural diagrams of the ProSliceFinder and ExCapNet CNN models (a). Accompanying the diagrams are tables that detail patient demographics, specifically Age and ISUP grade group, alongside the distribution of image data organized by ECE status based on post-surgery pathology results (b).

The model architecture is a CNN tailored for efficient image classification, utilizing depthwise separable convolutions to reduce computational complexity while maintaining performance. The network comprises three primary depthwise separable convolution blocks, each followed by batch normalization and ReLU activation, with subsequent downsampling via max pooling. The first block begins with a convolutional layer configured to retain the number of channels (3 in, 3 out), followed by a pointwise convolution that expands the channels to 12. This pattern of channel management and spatial reduction is mirrored in subsequent blocks with channels doubling from the output of the previous block (12 to 24, then 24 to 36). Post convolution operations, the spatial resolution is systematically reduced by half through max pooling, culminating in a substantial reduction of the input’s dimensionality. After the final pooling layer, the network transitions to a dense segment consisting of two fully connected layers separated by a dropout layer set at 60% to mitigate overfitting. The flattened output from the pooling layers is passed through these layers, with the final layer outputting a probability distribution across the predefined number of classes (2 in this case). The entire model is designed for compatibility with CUDA-enabled devices to leverage GPU acceleration, enhancing computational efficiency (**Figure 5a**).

The training of the ExCapNet model is facilitated by a robust preprocessing pipeline using the torchvision library’s transforms module. Image inputs are resized to 512x512 pixels to enhance detail recognition, randomly flipped horizontally to augment the dataset and improve generalization, and normalized to scale pixel values between -1 and 1. The training, validation, and test datasets are loaded using PyTorch’s DataLoader, with shuffled batches of 64 and 32 images for training and validation, respectively, to promote model robustness. The optimization process employs an RMSprop optimizer configured with a slightly elevated initial learning rate of 0.00001, an alpha of 0.9 for smoothing, and momentum set at 0.5 to accelerate convergence in the appropriate direction of the gradients. Weight decay is also used to reduce overfitting by penalizing large weights. The loss function employed is the CrossEntropyLoss, which is particularly effective for classification tasks involving multiple classes by combining LogSoftmax and NLLLoss in a single criterion.

## Data Availability

All data produced in the present study are available upon reasonable request to the authors. Clinical information such as pathology and age can be accessed through our GitHub repository at https://github.com/PKhosravi-CityTech/AutoRadAI. The MR images analyzed in this study are not publicly available due to privacy and security concerns and the sensitivity of medical data. These datasets are proprietary to the contributing institutions and are only available to researchers involved in institutional review board (IRB)-approved research collaborations with these centers.

## Data Distribution

As depicted in **Figure 5b**, the distribution of data across training, validation, and testing sets is meticulously balanced concerning patient age, preoperative biopsy pathological grade, and ECE outcomes. This figure provides a detailed overview of the dataset, which comprises a total of 4200 images from 701 patients allocated for training purposes, 1340 images from 200 patients designated for validation, and 622 images from 100 patients intended for testing.

Importantly, it highlights the mean age of patients within each subset—64.0 years for the training group, 65.41 years for the validation group, and 63.03 years for the testing group—thereby showcasing a consistent age distribution across all datasets. Additionally, a visual depiction of age distribution curves for each dataset set is presented, emphasizing the homogeneity of this demographic factor within our study. The figure’s lower panel elucidates the distribution of ECE status according to biopsy grade groups via a mosaic plot, thus confirming the balanced representation of various pathological grades and ECE outcomes across the study cohorts. The inclusion of preoperative biopsy grade and patient age as clinical parameters aims to enhance the predictive accuracy of a decision tree model, enabling a multimodal approach to estimating the likelihood of ECE.

### Statistical Tests and ML Predictive Modeling

To explore the relationship between patient age and preoperative International Society of Urological Pathology (ISUP) grade group, we utilized a blend of ML and DL approaches. Our ML toolkit comprised Decision Tree Classifier (DTC), Random Forest, and XGBoost algorithms, implemented via the Scikit-learn library in Python (version 3.10.12). Complementing these, we developed DL models using the PyTorch framework (version 1.9.0), to harness advanced computational techniques.

Before proceeding with the analysis, we prepared the dataset by converting categorical variables into dummy variables. This transformation, alongside the conversion of the ECE results into a binary outcome, allowed for seamless integration and analysis of both categorical and continuous variables. Our DL models underwent training and evaluation on a T4 GPU, chosen for its efficiency and capability to handle complex computations inherent in DL tasks.

The data was partitioned into a training set and a testing set following a 70/30 ratio to ensure a balanced representation of outcomes. The models were configured with a maximum depth of four to maintain generalizability and prevent overfitting. The decision structure export displays nodes rounded for clarity, with the tree proportionally colored to reflect class distribution at each decision juncture.

Comprehensive statistical analyses were conducted, including Specificity, Sensitivity, and Accuracy (ACC) evaluations, along with Receiver Operating Characteristic (ROC) curve analysis to calculate the Areas Under the Curve (AUCs). Comparisons against radiologists with varying expertise levels in prostate mpMRI readings were made using the Jaccard index and Cohen’s Kappa.

## Data availability

This study was approved by the AdventHealth Institutional Review Board (IRB), Orlando, FL, under protocol number 3009855250. The MR images analyzed in this study are not publicly available due to privacy and security concerns and the sensitivity of medical data. These datasets are proprietary to the contributing institutions and are only available to researchers involved in institutional review board (IRB)-approved research collaborations with these centers. However, clinical information such as pathology and age can be accessed through our GitHub repository. The methodologies developed for AutoRadAI are not specific to the datasets used in this study, allowing researchers to apply our deep-learning models to other relevant MR imaging and clinical data to assess their effectiveness in different contexts.

## Code availability

In our commitment to transparency and collaborative advancement, The official source code repository for AutoRadAI is publicly accessible on GitHub at https://github.com/PKhosravi-CityTech/AutoRadAI. In addition, AutoRadAI is available through a web-based user interface at https://autoradai.anvil.app designed to allow clinicians and researchers to explore its functionality.

### Acknowledgments

This research was partially supported by Barrel Aged Charities, a 501(c)(3) charitable organization. Further information is available at BarrelAgedCharities.org.

## Contributions

P.K., S.S., S.S.V., and V.P. conceived the study. P.K. developed the methodology, designed the algorithmic techniques, and developed the entire pipeline. S.S. and M.B. generated the datasets and prepared and labeled images according to various grade groups. S.M. contributed to data processing and result visualization. A.A. created the web interface, while D.M.D. and C.S. manually scored the MR images for radiology scores in the test set, serving as the human benchmark to compete with the AI’s performance. M.C.M., I.H., and A.J.H. provided critical reviews of the manuscript, contributing comments that improved the methodology. P.K. drafted the manuscript, and all authors participated in reviewing, editing, and approving the final version.

